# Machine learning reduced gene/non-coding RNA features that classify Schizophrenia patients accurately and highlight insightful gene clusters

**DOI:** 10.1101/2020.06.08.20125906

**Authors:** Yichuan Liu, Hui-Qi Qu, Xiao Chang, Lifeng Tian, Joseph Glessner, Patrick A. M. Sleiman, Hakon Hakonarson

## Abstract

Schizophrenia (SCZ) is a chronic and severely disabling neurodevelopmental disorder that affects people worldwide. RNA-seq has been a powerful method to detect the differentially expressed genes/non-coding RNAs in patients; however, due to overfitting problems differentially expressed targets (DETs) cannot be used properly as biomarkers. In this study, dorsolateral prefrontal cortex (dlpfc) RNA-seq data from 254 individuals’ was obtained from the CommonMind consortium and analyzed with machine learning methods, including random forest, forward feature selection (ffs), and factor analysis, to reduce the numbers of gene/non-coding RNA feature vectors to overcome overfitting problem and explore involved functional clusters. In 2-fold shuffle testing, the average predictive accuracy for SCZ patients was 67% based on coding genes, and the 96% based on long non-coding RNAs (lncRNAs). Coding genes were further clustered into 14 factors and lncRNAs were clustered into 45 factors to represent the underlying features. The largest contribution factor for coding genes contains number of genes critical in neurodevelopment and previously reported in relation with various brain disorders. Genomic loci of lncRNAs were more insightful, enriched for genes critical in synapse function (p=7.3E-3), cell junction (p=0.017), neuron differentiation (p=8.3E-3), phosphorylation (8.2E-4), and involving the Wnt signaling pathway (p=0.029). Taken together, machine learning is a powerful algorithm to reduce functional biomarkers in SCZ patients. The lncRNAs capture the characteristics of SCZ tissue more accurately than mRNA as the formers regulate every level of gene expression, not limited to mRNA levels.

## Introduction

Schizophrenia (SCZ) is a complex biological disorder involves combined effect of many genes, each conferring a small increase in susceptibility to the illness^1^. The redundancy of the gene networks underlying SCZ indicates that many gene combinations have the potential to result in a brain dysfunction that can manifest as SCZ or a related neurodevelopmental disorder^2^. Next generation sequencing (NGS) enables measures of the transcriptome gene expression through RNA-seq, however, expressed genes cannot be used as biomarkers in many diseases that involve complex genetic networks, due to high noise level from large number of genes and small number of samples. While the current solution is considering only differentially expressed targets (DETs) between SCZ and healthy controls, this often has multiple potential problems. For example, single or a small number of differentially expressed genes may not be clinically important for SCZ, indicating that a more comprehensive analysis are necessary to reveal underling genetic network for SCZ. Selection criteria for DETs are arbitrary, and while many researchers are using adjusted p value of 0.05 as a cut-off, this static standard often brings more ambiguity and neglects downstream analysis. Even if only DETs were selected with a p value cut-off as features for labeling or prediction, the DET number could still be too large which means that an overfitting problem could exists theoretically. The key to address overfitting from too many features is to effectively reducing the number of features.

Beside coding genes, non-coding RNAs, especially long non-coding RNAs (lncRNAs) are important factors in shaping SCZ networks and are dynamically regulated by neuronal activation^3^, and should therefore also be considered as potential feature vectors for SCZ gene network regulations. In this study, we acquired dorsolateral prefrontal cortex (dlpfc) samples’ RNA-seq data from 254 subjects from the CommonMind consortium (120 SCZ patients and 130 healthy controls, all non-Hispanic Caucasian). We then applied machine learning algorithms, including random forest, forward feature selection (ffs), and factor analysis to reduce the number of expressed genes into small list of feature vectors, in order to solve the overfitting problem. Two-fold shuffle tests showed that these selected feature vectors could accurately label SCZ patients versus controls. Selected genes were further clustered into gene modules through factor analysis, to explore potential functional units within the complex underlying genetic networks in SCZ.

## Methods & Materials

### RNA-seq data for dorsolateral prefrontal cortex (dlpfc) samples

RNA-seq data of dorsolateral prefrontal cortex (dlpfc) samples were obtained from CommonMind consortium FTP sites directly. In order to mimic other factors, we only selected SCZ patients and controls who are of European ancestry (EA). A total of 254 RNA-seq BAM files were obtained include 120 SCZ patients and 134 healthy controls. Samples with a minimum of 50 million mapped reads and less than 5% rRNA-aligned reads were retained for downstream analysis. Based on the consortium’s description, RNA was isolated from 50□mg homogenized tissue in Trizol using the RNeasy kit based on the instructional protocol. The mean total RNA yield was 15.3□µg (±5.7). The RNA integrity number (RIN) was determined by fractionating RNA samples on the 6000 Nano chip (Agilent Technologies) on the Agilent 2100 Bioanalyzer. The mean RIN was 7.7 (±0.9), and the mean ratio of 260/280 was 2.0 (±0.02). Processing order was re-randomized prior to ribosomal RNA (rRNA) depletion. Briefly, rRNA was depleted from about 1□µg of total RNA using Ribo-Zero Magnetic Gold kit (Illumina/Epicenter Cat # MRZG12324) to enrich for polyadenylated coding RNA and non-coding RNA. The sequencing library was prepared using the TruSeq RNA Sample Preparation Kit v2 (RS-122–2001-48 reactions) in batches of 24 samples. A pool of 10 barcoded libraries was layered on a random selection of two of the eight lanes of the Illumina flow cell bridge amplified to ∼250 million raw clusters. One-hundred base pair paired end reads were obtained on a HiSeq 2500. The sequence data were processed for primary analysis to generate QC values.

### Gene/non-coding RNAs expression matrix

The genomic template used for coding genes expressions is hg19 refSeq, and long non-coding RNAs template is GENCODE version 19^4^. The expression matrix was generated based on Cuffnorm functions in Cufflink package version 2.2.1^5^, and the SCZ and controls groups are normalized. To eliminate potential noisy signals, the gene expression FPKM value less than 2 and genes/lncRNAs with collinearity over 80% were removed.

### Gene reductions using machine learning algorithms

Multiple machine learning algorithms include random forest, forward feature selection (ffs), and factor analysis, were applied to select and reducing the informative gene/lncRNA features between SCZ and controls. Random forest is one of the most widely used algorithms for feature selection, which computes relative importance or contribution of each gene feature in the prediction, then scales the relevance down so that the sum of all scores is 1. All the genes/lncRNAs with zero relative importance were removed. Second algorithm forward feature selection (ffs) is one of the most common method to reduce number of features for machine learning inputs by trying to find the best features which improve the performance of the model. The modeling codes are based on based on the Scikit-learn package in Python language^6^.

In order to test the predictive abilities for selected gene/lncRNA features, we applied a 2-fold shuffle testing. In other words, the SCZ and control samples were split into 1:1 ratio for 50 rounds randomly, one set used as training data and another one used as independent testing set. Gene/lncRNA features were selected as described in previous paragraph for training data (to overcome overfitting problem, the only parameter altered is we required random forest relative importance > 0.0005 rather than > 0), then a random forest classifier is applied to label whether the sample is SCZ or control in testing data based on training data gene/lncRNA features.

Factor analysis was applied to entire sample set for further clustering gene/lncRNA features. Factor analysis is a statistical method used to describe variability among observed, correlated variables in terms of a potentially lower number of unobserved variables called factors, and the methods have been proven to be a good interpreter for gene networks and pathways. The Python-based factor_analyzer package was used in the analysis.

## Results

The statistical analysis and fold changes of genes were calculated. Altogether, 10,100 genes showed nominal significance with P<0.05. Among the 10,100, expression of 3,483 genes were down-regulated, and expression of 6,617 genes were up-regulated. Using the WebGestalt (WEB-based Gene SeT AnaLysis Toolkit) web tool^7^, over-representation analysis (ORA) by the Reactome approach^8^ highlighted genes involved in mitochondrial function as down-regulated; and genes involved in gene transcription as upregulated (Supplementary Table 1 and 2).

**Table 1.**
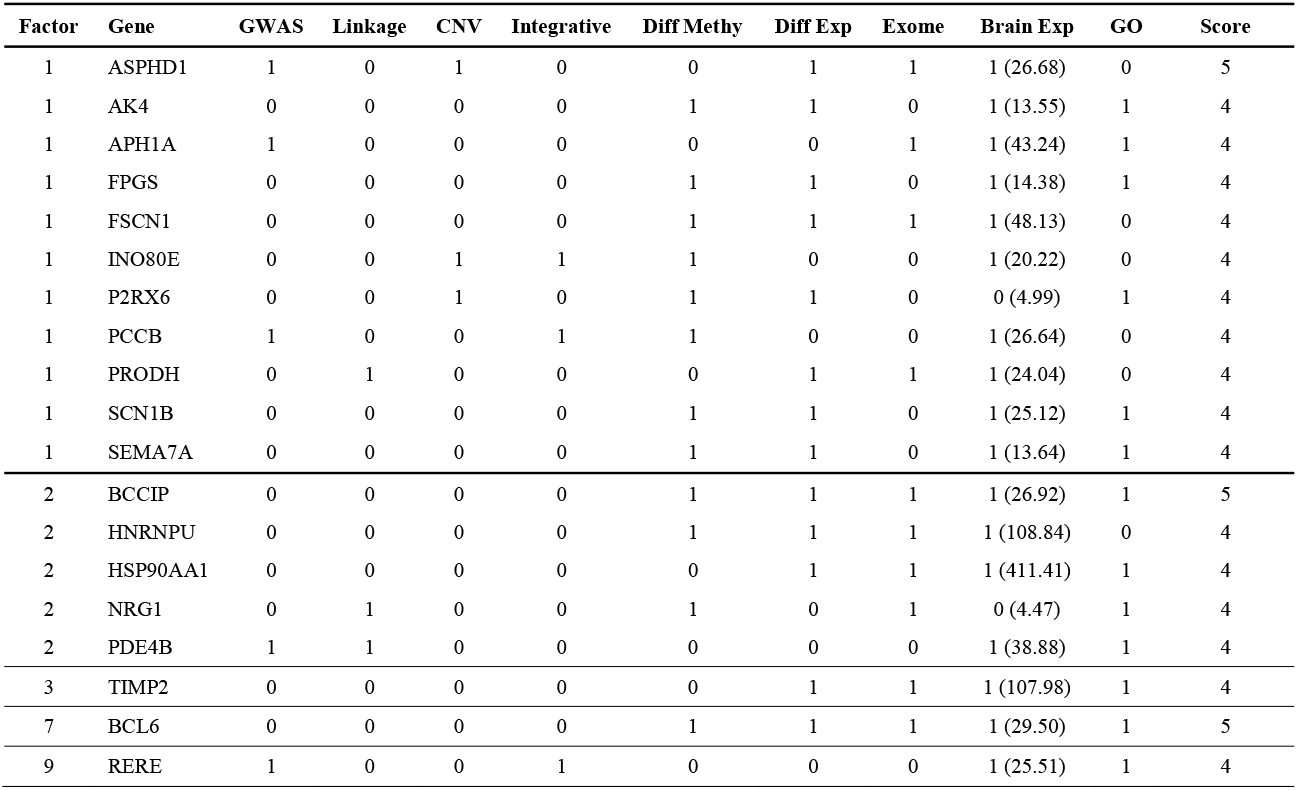
Genes in gene factors have high supportive evidences

**Table 2.** Target genes in lncRNA factors have high supportive evidences

| Factor | lncRNA | target Gene | GWAS | Linkage | CNV | Integrative | Diff Methy | Diff Exp | Exome | Brain Exp | GO | Score |
| --- | --- | --- | --- | --- | --- | --- | --- | --- | --- | --- | --- | --- |
| 1 | ENSG00000247735.2 | ASPHD1 | 1 | 0 | 1 | 0 | 0 | 1 | 1 | 1 (26.68) | 0 | 5 |
| 1 | ENSG00000232912.1 | RERE | 1 | 0 | 0 | 1 | 0 | 0 | 0 | 1 (25.51) | 1 | 4 |
| 1 | ENSG00000235770.1 | FN1 | 0 | 1 | 0 | 0 | 0 | 0 | 1 | 1 (38.33) | 1 | 4 |
| 1 | ENSG00000235831.2 | ITPR1 | 0 | 0 | 0 | 0 | 1 | 0 | 1 | 1 (25.59) | 1 | 4 |
| 1 | ENSG00000239569.2 | SRPK2 | 1 | 0 | 0 | 0 | 1 | 0 | 0 | 1 (75.17) | 1 | 4 |
| 1 | ENSG00000243762.1 | RANBP1 | 0 | 1 | 1 | 0 | 1 | 0 | 0 | 1 (30.73) | 0 | 4 |
| 1 | ENSG00000247735.2 | SEZ6L2 | 1 | 0 | 1 | 0 | 0 | 0 | 0 | 1 (38.77) | 1 | 4 |
| 1 | ENSG00000257126.1 | FOXG1 | 1 | 0 | 0 | 0 | 1 | 0 | 0 | 1 (22.05) | 1 | 4 |
| 1 | ENSG00000261220.2 | ST3GAL1 | 0 | 0 | 0 | 0 | 1 | 0 | 1 | 1 (8.93) | 1 | 4 |
| 1 | ENSG00000271849.1 | PJA2 | 0 | 0 | 0 | 0 | 1 | 0 | 1 | 1 (144.54) | 1 | 4 |
| 2 | ENSG00000224563.1 | BCL6 | 0 | 0 | 0 | 0 | 1 | 1 | 1 | 1 (29.50) | 1 | 5 |
| 2 | ENSG00000226978.1 | MAGI2 | 0 | 1 | 0 | 0 | 1 | 0 | 0 | 1 (24.46) | 1 | 4 |
| 2 | ENSG00000236031.1 | AKT3 | 1 | 0 | 0 | 0 | 0 | 0 | 1 | 1 (35.93) | 1 | 4 |
| 2 | ENSG00000248816.1 | TENM3 | 0 | 0 | 0 | 0 | 1 | 1 | 0 | 1 (8.06) | 1 | 4 |
| 2 | ENSG00000272367.1 | RASA1 | 0 | 0 | 0 | 0 | 1 | 1 | 0 | 1 (20.45) | 1 | 4 |
| 3 | ENSG00000272989.1 | DLG1 | 0 | 0 | 1 | 0 | 0 | 1 | 1 | 1 (52.38) | 1 | 5 |
| 3 | ENSG00000239569.2 | SRPK2 | 1 | 0 | 0 | 0 | 1 | 0 | 0 | 1 (75.17) | 1 | 4 |
| 3 | ENSG00000273164.1 | PRODH | 0 | 1 | 0 | 0 | 0 | 1 | 1 | 1 (24.04) | 0 | 4 |
| 3 | ENSG00000273164.1 | DGCR2 | 0 | 1 | 1 | 0 | 0 | 0 | 1 | 1 (30.80) | 0 | 4 |
| 37 | ENSG00000248816.1 | TENM3 | 0 | 0 | 0 | 0 | 1 | 1 | 0 | 1 (8.06) | 1 | 4 |

### Accuracy measure for labeling SCZ patients based on 2-fold shuffle testing

As described in the method section, 2-fold shuffle testing was applied to test the labeling prediction 50 times. Reduced genes, based on multiple machine learning methods, ranged from 36 to 282, showed certain level of accuracy (∼67%) in classifying SCZ patient’s dorsolateral prefrontal cortex (dlpfc) samples versus healthy controls (Figure 1a). In contrast, machine learning methods reduced long non-coding RNAs (lncRNAs), ranging from 32 to 110 lncRNAs, and showing extremely high accuracy in classifying SCZ patients (accuracy level ∼99%) (Figure 1b). The results indicate the essential regulation network for SCZ brain tissues are very stable although the expressed gene network may diversify more across individuals.

**Figure 1.**
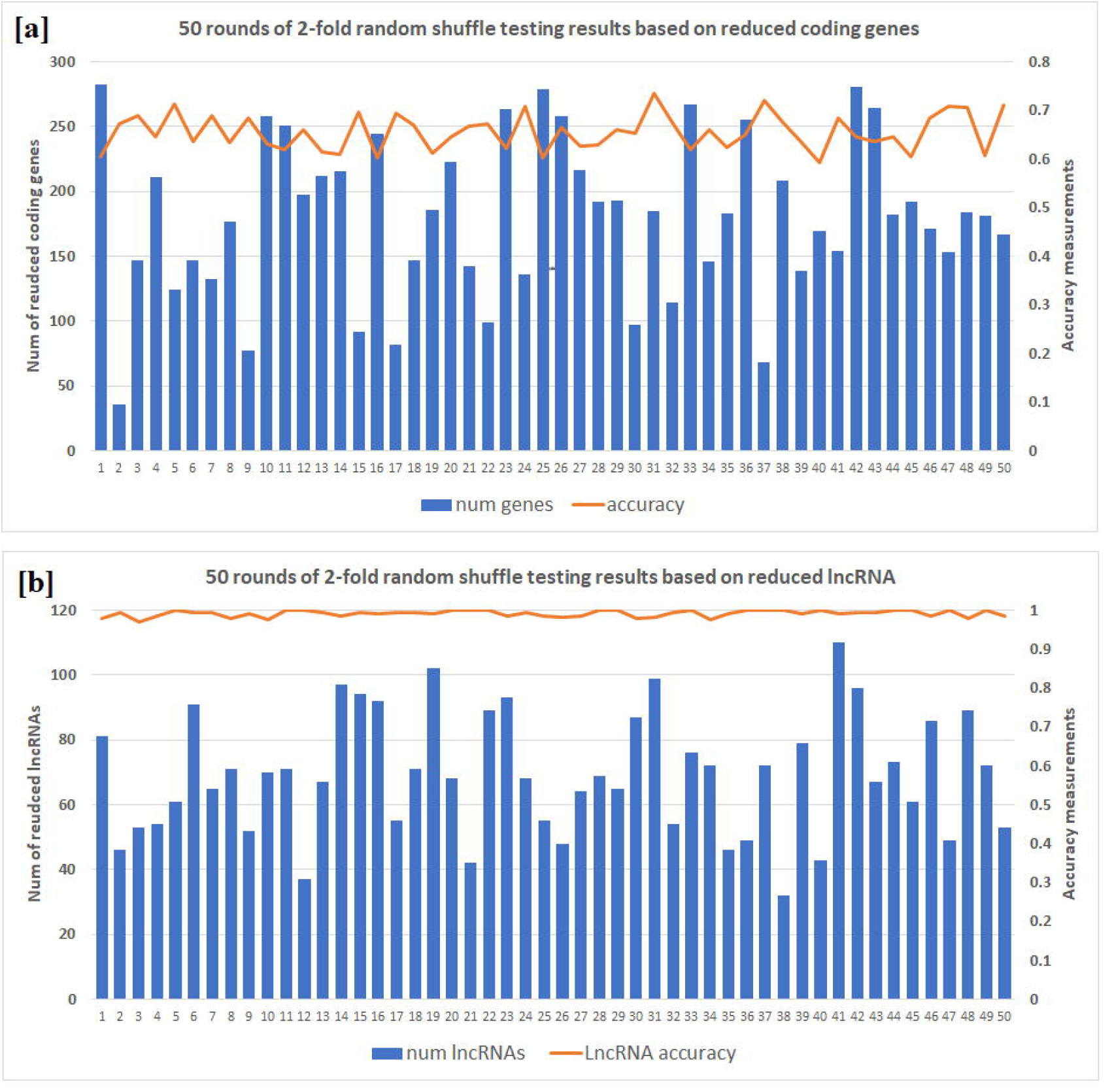
2-fold random shuffle testing results for 50 rounds. X-axis is the round number, Y_1 axis (left) is the number of reduced genes [a] and lncRNAs [b], Y_2 axis (right) is the accuracy measurement ranged from 0 to 1.

### Selected Gene/lncRNA feature based on Machine learning algorithm

After multiple layers of filtering, including the machine learning methods, the number of genes reduced from 27,101 to 734 in the total of 254 dorsolateral prefrontal cortex (dlpfc) samples (Figure 2a). Expressions of all these genes have nominally statistical significance with P value range from 1.15E-09 to 6.45E-03. Among the 734 genes (Supplementary Table 3), 412 were downregulated; and 322 genes were upregulated. These genes found to be enriched in glycosaminoglycan metabolic processes (adjusted p value = 3.1E-3), aminoglycan metabolic processes (adjusted p value = 3.7E-3), mucopolysaccharide metabolic processes (adjusted p value = 1.2E-2), based on Gene Ontology^9^. A total of 13,871 long non-coding RNAs (lncRNAs) identified in GENCODE were reduced to 605 using comparable pipelines. The results suggest that by combining multiple machine learning methods is a powerful tool to reduce the number of gene features which represent the variations of the data.

**Figure 2.**
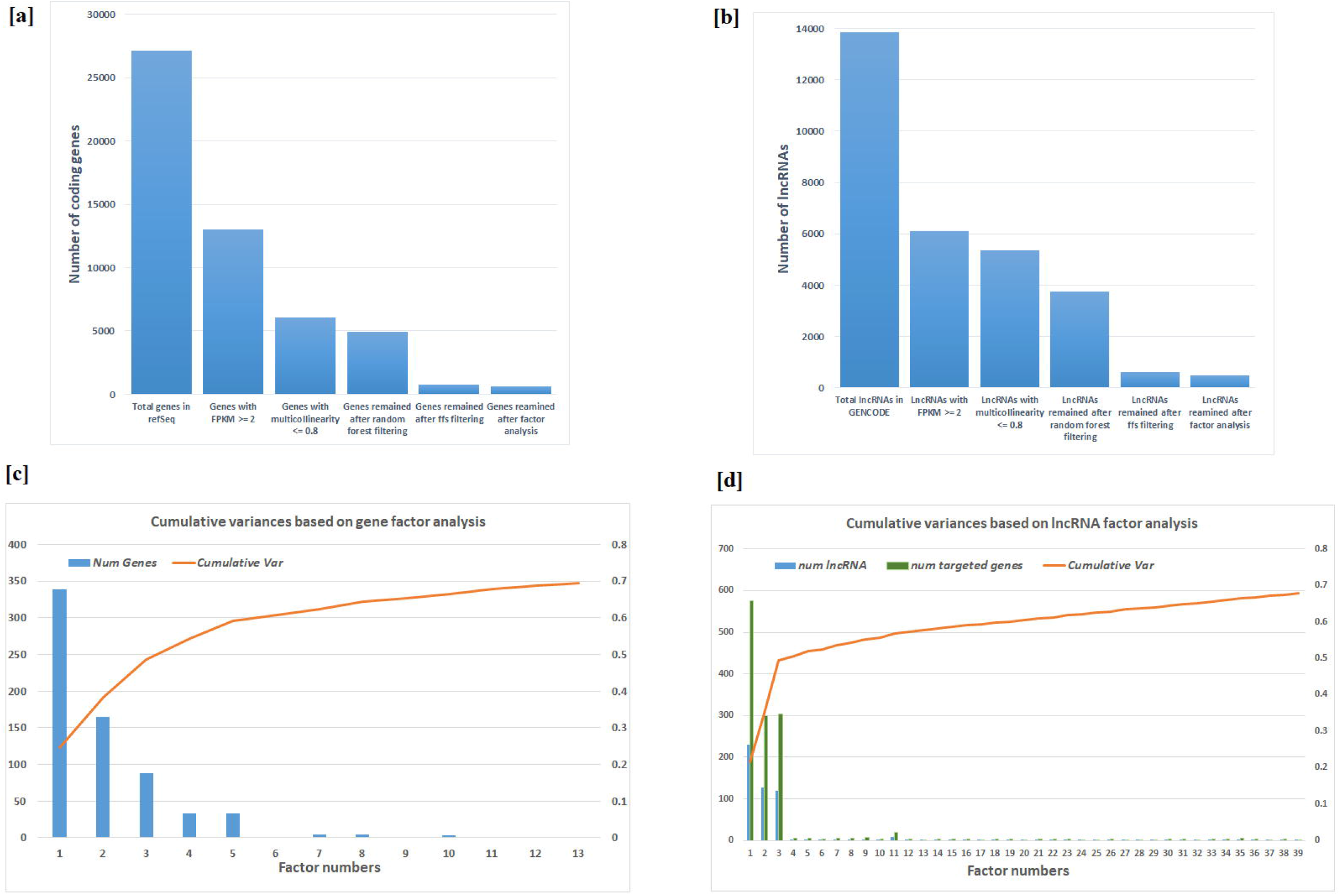
Number of genes after filtering and factor analysis cumulative curve. [a] number of feature vectors for coding genes after multiple filtering methods; [b] number of feature vectors for lncRNAs after multiple filtering methods; [c] factor analysis cumulative curve and number of remain coding-gene feature vectors; [d] factor analysis cumulative curve and number of remain lncRNAs and their targeted genes feature vectors

### Factors and potential gene modules from factor analysis

The 734 genes were clustered into 66 factors, where the first 13 factors contribute ∼70% of the variances (Figure 2c, Supplementary Table 4). Factor 1 contains 339 genes and contributes 24.5% of the total variance, all genes in factor 1 are down regulated in SCZ versus health controls, as shown by differential expression in CommonMind consortium database^10^. Not unexpectedly, over-representation analysis (ORA) by the Human Phenotype Ontology (HPO) shows that factor 1, as the major factor of SCZ transcriptome, contains a number of genes critical in neurodevelopment that are involved in various brain disorders, including HP:0001298_Encephalopathy, HP:0001098_Abnormal fundus morphology, and HP:0004329_Abnormal morphology of the posterior segment of the globe. Enrichment analysis by biological pathways (Figure 4a) highlighted sulfur compound biosynthetic processes (adjusted p value = 4.8E-4), glycosaminoglycan metabolic processes (adjusted p value = 8.1E-3), and glycoprotein metabolic processes (adjusted p value = 0.019). Eight genes were identified in previous PG2 GWAS studies^11^, including *APH1A* (rs140505938, p value = 4.49E-10), *ASPHD1* (rs11646127, p value = 4.55E-11), *BRINP2* (rs6670165, p value = 4.45E-08), *CHRM4* (rs7951870, p value = 1.26E-11), *INO80E* (rs11646127, p value = 4.55E-11), *PCCB* (rs7432375, p value = 7.26E-11), *SPCS1* (rs3617, p value = 4.26E-11), and *TAC3* (rs61937595, p value = 2.02E-12), all these hotspots were also identified in CLOZUKe^12^. Of note, 159 out of the 339 genes (46.9%) have at least one supportive evidence from previous knowledge, and 12 genes have at least four supportive evidences (Table 1).

**Figure 3.**
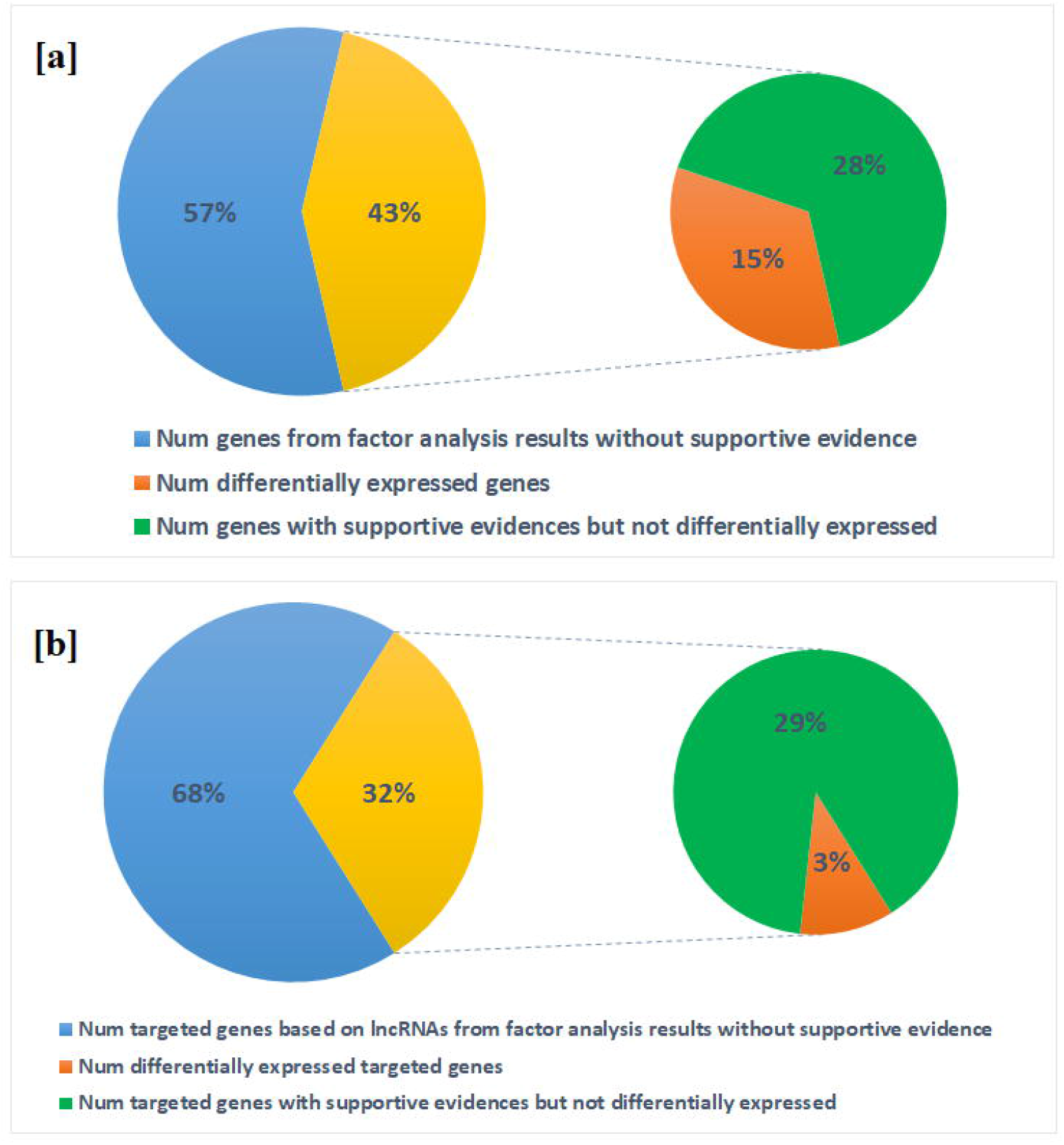
Portion of differentially expressed genes/lncRNAs versus genes/lncRNAs with at least one supportive evidence: [a] coding genes after factor analysis; [b] targeted coding genes based on lncRNA genomic locus.

**Figure 4.**
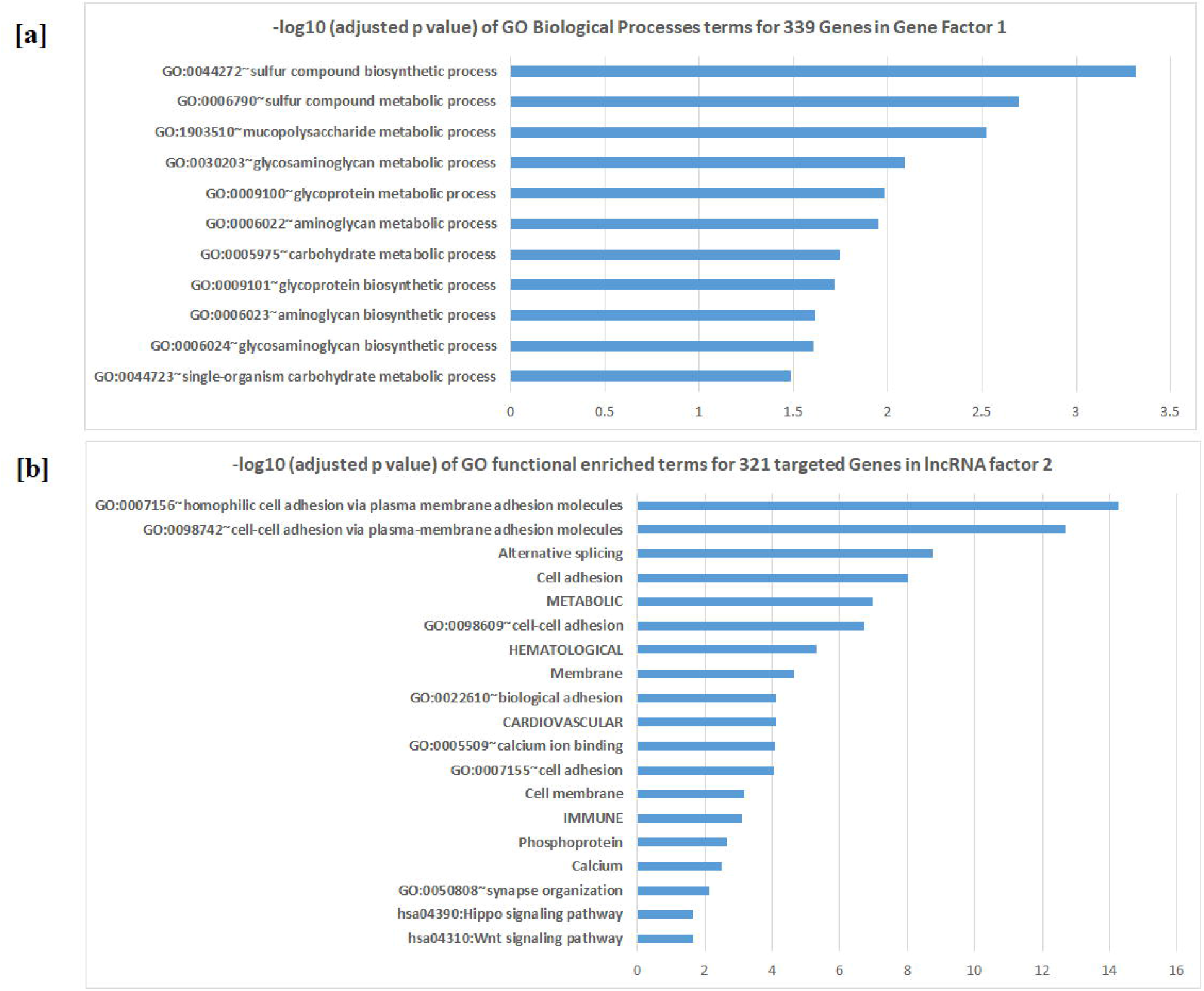
-log10 (Adjusted p value) scale for Enriched functional pathways: [a] Gene factor 1; [b] Targeted genes in lncRNA factor 2

A total of 166 genes clustered into factor 2 and majority of them (94%) are up regulated in SCZ. Factor 2 contributes 13.9% of the data variance (Figure 2c, Supplementary Table 4), 11 genes (6.7%) were found to be differentially expressed and none of them are down regulated in SCZ. A total of 65 out of 165 genes (39.3%) have at least one supportive evidence from previous knowledge, and 5 genes have at least four supportive evidences (Table 1). Enrichment analysis by biological pathways highlighted phosphorylation (adjusted p value = 5.1E-3). There are 88, 33, and 33 genes in factor 3, 4 and 5 (Supplementary Table 4), respectively, contributing 10%, 5.7% and 4.8% of the variance, respectively.

For lncRNAs, factor analysis resulted in 91 factors, where the first 45 factors (contain 496 lncRNAs) explain ∼70% of the variations of the expression data (Figure 2d, Supplementary Table 5). The genes closest to the lncRNA binding sites were identified as their target genes, therefore in order to explore the full potential targets, the closest three genes were selected. A total of 1193 genes were selected and found to be enriched in multiple disease category^13^, METABOLIC (adjusted p value = 2.8E-5), CARDIOVASCULAR (3.2E-2), HEMATOLOGICAL (7.3E-3), IMMUNE (4.8E-2), and post translational modifications such as phosphorylation (8.2E-4). Multiple neurodevelopmental related pathways, much more informative than the above mRNA pathway analysis, are highlighted for the lncRNAs’ targeted gene sets, including regulation of transferase activity (adjusted p value =1.8E-3), neuron projection morphogenesis (adjusted p value = 4.8E-3), positive regulation of nervous system development (adjusted p value = 4.6E-3); regulation of kinase activity (adjusted p value = 5.7E-3), neuron differentiation (adjusted p value = 8.3E-3), and neuron projection development (adjusted p value =8.8E-3).

## Discussion

Growing evidence indicates that distinct neuronal ncRNAs, particularly lncRNAs, are likely to influence the development of neurodevelopmental diseases, including SCZ^3^. However, to date, neither genes in neurodevelopmental networks nor the lncRNAs in regulation processes have been successfully applied as feature vectors to label the phenotypic status of the patients. The main obstacle is the number of genes/lncRNAs as predictive features is huge (thinking about over twenty thousand genes and ten thousand lncRNAs), and the number of biological samples, especially brain tissues, is usually small due to the difficulty in sample collections. Therefore, predictive models are deemed to fail due to overfitting issues. Currently, the solution for this problem is to select a small number of genes that are differentially expressed between SCZ and controls. While this may help, the algorithm and criteria used for selecting these gene remains controversial, and genes contributing to the network with small effects and less statistical significance may be missed.

Machine learning methods have been proven to be effective in reducing the feature vectors while capturing essential data differences in studies of many fields, include genetic expression studies ^14,15^. In this study, we applied multiple machine learning layers for expressed genes in dorsolateral prefrontal cortex (dlpfc) RNA-seq data from 254 samples from the CommonMind consortium, in order to show that reduced gene/lncRNA features could accurately classify SCZ patients versus healthy controls. Combining machine learning methods, such as random forest and forward feature selection (ffs) for expressed genes/lncRNAs, the number of genes was significantly reduced from over twenty-five thousand to averagely ∼180 genes while the lncRNA number was reduced from thirteen thousand to averagely ∼70 through the simulations. The 2-fold shuffle tests (samples split to 1:1 ratio, half used as training data and rests used as testing data) applied in 50 separate rounds (Figure 1) shows that reduced gene feature vector has modest power (∼67% accuracy) in classifying SCZ patients versus healthy controls, whereas lncRNAs could serve as an effective predictor (∼99% accuracy). These results demonstrate that machine learning has the potential to be an alternative methods to detect the essential differences of gene expressions in SCZ, also the regulation networks involved in lncRNAs are more stable than gene expression networks, in other words, gene expressions remain highlydiverse for different persons but the lncRNAs expression seems universal among different individuals. More importantly, as demonstrated by our findings that the average predictive accuracy for SCZ patients is 67% based on coding genes, and 96% based on lncRNAs, lncRNAs represent a more accurate biomarker for the SCZ transcriptome. The lncRNAs regulates every level of gene expression, including but not limited to mRNA levels, which may explain why lncRNAs capture the characteristics of SCZ tissue more accurately^16^.

Furthermore, the reduced gene/lncRNA features were clustered based on factor analysis to form the gene modules. Our study showed that major factors were enriched for genes important in neurodevelopment and brain disorders, thereby proving the validity of the dimension reduction process. While genes in major factors show enrichment of SCZ related pathways or neurodevelopmental associated network and can serve as a proof-of-principle of this study, other factors may harbor novel knowledge about SCZ and warrant further study. Genes within each factor have higher portion of SCZ supportive evidences while known differentially expressed genes only counted small portion of genes in each factor (Figure 3). Combining these clues together indicates that the machine learning models capture contributing genes more effectively compared to traditional differential expression tests. SCZ genetics involves combined effect of many genes, each conferring a small increase in susceptibility to the illness^17^.

Genes in certain factors highlighted SCZ associated networks and the biochemical molecules, synthesis/metabolism, as neuro system modulators. For example, 339 genes in gene factor 1 were enriched in sulfur compound biosynthetic and metabolic processes (adjusted p value = 4.8E-3). Sulfur is an essential chemical for proteinogenic amino acid methionine (Met), and methionine-folate cycle-dependent one-carbon metabolism is implicated in the pathophysiology of SCZ while deficiencies in the one-carbon metabolism components folate is consistent findings in SCZ patients^18^. These genes are also enriched in glycosaminoglycan metabolic processes (adjusted p value = 8.1E-3), glycosaminoglycan and neurotransmitter metabolism have been reported to associated with SCZ in cerebral cortex^19^. Other enriched chemical synthesis procedures for genes in factor 1 include glycoprotein metabolic processes (adjusted p value = 0.019) where P-glycoprotein a major efflux pump in the blood-brain barrier, has a profound effect on entry of drugs, peptides and other substances into the central nervous system^20^. For lncRNA factor 2, 321 targeted genes (union of 128 lncRNAs’ top three closest genes) were significantly enriched in cell adhesion (adjusted p value = 9.7E-9), phosphorylation (adjusted p value = 2.2E-3), SCZ associated pathways include calcium ion binding^21^ (adjusted p value = 8.1E-5) as well as the Wnt signaling pathway^22^ (adjusted p value = 0.022) (Figure 4b). In contrasts, many factors although with high consistent regulation tendency have no known genetic functions. For example, gene factor 2 (94% genes up regulated in SCZ, 39.3% have at least one supportive evidences, and only 11% of the genes are known to differentially expressed in SCZ), gene factor 3 (78% genes up regulated in SCZ, 31.8% have at least one supportive evidences, and only 4.5% of the genes are known to differentially expressed), gene factor 4 (70% genes down regulated in SCZ, 42.4% have at least one supportive evidences, and only 15% of the genes are known to differentially expressed), and lncRNA factor 1, which contains 229 lncRNAs (597 target genes) and 10 target genes with high supportive evidences (score >= 4, Table 2) but no enrichment were identified for biological functions. Taken together, these factors are potential targets for researchers to explore in further studies.

## Data Availability

All data generated or analysed during this study are included in this published article.

## Acknowledgments

DLPFC data were generated as part of the CommonMind Consortium supported by funding from Takeda Pharmaceuticals Company Limited, F. Hoffman-La Roche Ltd, and from NIH grants R01MH085542, R01MH093725, P50MH066392, P50MH080405, R01MH097276, RO1-MH-075916, P50M096891, P50MH084053S1, R37MH057881, and R37MH057881S1, HHSN271201300031C, AG02219, AG05138, and MH06692. Brain tissue for the study was obtained from the following brain bank collections: the Mount Sinai NIH Brain and Tissue Repository, the University of Pennsylvania Alzheimer’s Disease Core Center, the University of Pittsburgh NeuroBioBank and Brain and Tissue Repositories, and the NIMH Human Brain Collection Core. CMC Leadership: Pamela Sklar, Joseph Buxbaum (Icahn School of Medicine at Mount Sinai), Bernie Devlin, David Lewis (University of Pittsburgh), Raquel Gur, Chang-Gyu Hahn (University of Pennsylvania), Keisuke Hirai, Hiroyoshi Toyoshiba (Takeda Pharmaceuticals Company Limited), Enrico Domenici, Laurent Essioux (F. Hoffman-La Roche Ltd), Lara Mangravite, Mette Peters (Sage Bionetworks), Thomas Lehner, and Barbara Lipska (NIMH).

## Ethical approval

This study had been approved by the Children’s Hospital of Philadelphia. All the patients who participated in this project have been consented and they agree to publish the results.

## Competing interests

The authors declare that they have no competing financial interests.

## Notes

### Competing Interest Statement

The authors have declared no competing interest.

